# Pharmacist Interventions and Barriers to Pharmaceutical Care in a Secondary-Care Facility in Nigeria

**DOI:** 10.64898/2026.01.23.26344744

**Authors:** Omolayo Umaru

## Abstract

**Background:** Pharmaceutical care is a patient-centered practice model in which pharmacists assume responsibility for identifying, preventing, and resolving medication-related problems to optimize therapeutic outcomes. In Nigeria, pharmaceutical care activities within hospital settings have expanded over time; however, evidence describing the frequency of specific pharmacist intervention activities and the structural barriers influencing their delivery at the facility level remains limited.

**Objectives:** This study aimed to (1) describe the types and frequency of pharmaceutical care intervention activities performed by pharmacists in a secondary-care facility in Nigeria, and (2) identify barriers limiting effective pharmaceutical care delivery in this setting.

**Methods:** A descriptive cross-sectional survey was conducted among pharmacists at a 50-bed secondary-care facility in Ibadan, Nigeria. All pharmacists employed at the facility were invited to participate. Data was collected using a structured, self-administered questionnaire assessing pharmacist demographics, frequency of intervention activities, perceived barriers to pharmaceutical care delivery, and recommendations for improvement. Quantitative data was analyzed descriptively using frequencies and percentages.

**Results:** Twelve pharmacists participated in this study (response rate: 92.3%). Pharmacists reported frequent engagement in clinically meaningful interventions, including: therapeutic substitutions (87%), provision of drug information to prescribers (83%), prevention of duplicate therapy, and dosage adjustments. Less frequently reported activities included adverse drug reaction reporting and counseling on medication storage. Major barriers to pharmaceutical care delivery were primarily system-related, including: limited access to patient medical records (92%), inadequate privacy for patient consultations (88%), and inefficient documentation systems (83%). Staffing levels and clinical knowledge were not perceived as major barriers.

**Conclusion:** Pharmacists in this secondary-care facility actively engaged in medication optimization and patient safety-oriented interventions; however, system-level constraints limited the delivery of comprehensive pharmaceutical care. Addressing infrastructural and documentation barriers is essential to strengthen pharmaceutical care practice in secondary-care hospital settings in Nigeria and similar low-and middle-income country (LMIC) contexts.

## INTRODUCTION

Pharmaceutical care is a patient-centered practice model in which pharmacists assume responsibility for identifying, preventing, and resolving medication-related problems to optimize therapeutic outcomes and improve patients’ quality of life (Hepler & Strand, 1990; Cipolle et al., 2012). In hospital settings, pharmacist’s roles - such as therapeutic substitution, dosage optimization, prevention of duplicate therapy, and provision of drug information to prescribers contribute to safer prescribing and improved medication use. The expansion of clinical pharmacy services has been associated with better prescribing practices, reduced medication errors, and enhanced collaboration within multidisciplinary health teams (Kaboli et al., 2006; Bond et al., 2002). In Nigeria, the adoption of pharmaceutical care and clinical pharmacy services has increased over time, but implementation remains uneven, particularly in facilities where limited infrastructure, fragmented information systems, and poor documentation practices may constrain pharmacists’ ability to deliver comprehensive services (Ogbonna et al., 2015). While prior literature has described pharmacists’ attitudes toward pharmaceutical care and general barriers to its implementation in Nigeria, these studies have largely focused on broad perceptions rather than practice-level realities (Oparah & Eferakeya, 2005; Akande-Sholabi & Akinbitan, 2022). Consequently, fewer studies have provided facility-level evidence that quantifies the frequency of specific pharmacist intervention activities while simultaneously examining the structural barriers that shape day-to-day practice in secondary-care hospital environments.

This study therefore aimed to (1) describe the types and frequency of pharmacist pharmaceutical care activities performed in a secondary-care facility in Ibadan, Nigeria, and (2) identify barriers perceived by pharmacists as limiting the effective delivery of pharmaceutical care in this setting.

## METHODS

### Study Design and Setting

Following Institutional Review Board approval, a descriptive cross-sectional survey was conducted at the University Health Services Department (Jaja Clinic), a 50-bed secondary-care facility located within the University of Ibadan, Nigeria. Formerly a primary-care center, the Clinic was upgraded to a secondary-care hospital in 2016; and provides medical services exclusively to students and staff of the university, running specialist clinics including the Hypertension, Diabetes, Asthma, Psychiatry, Maternity and Arthritis clinics. The Pharmacy unit, which handles outpatient prescriptions and patient counseling, was the focus for this study. The study was conducted within a 5-week period. Purposive sampling was used to recruit the pharmacists working at the facility. Only one pharmacist declined to participate.

### Study Participants

All 13 pharmacists employed at the facility - including full-time staff, interns, and National Youth Corps pharmacists, were invited to participate; and 12 of them completed and returned the questionnaire, representing a 92.3% response rate.

### Data Collection

A structured, self-administered questionnaire to assess pharmacists’ pharmaceutical care intervention activities and perceived barriers was developed, and pilot tested with two pharmacists in a different health system equivalent in size and operations. The questionnaire was further refined based on feedback from the pilot test. Additionally, two other pharmacists from the other health system served as expert reviewers, providing critical insights that further improved the questionnaire items.

The questionnaires included close-ended items with Likert-type scales or categorical response options, as well as open-ended items. These included (1) pharmacists’ demographics (multiple-choice questions), (2) types and frequency of intervention activities performed (Likert-scale), (3) barriers to effective pharmaceutical care delivery (Likert-scale), and (4) recommendations for improving pharmaceutical care services (open-ended questions).

Participants’ consent was obtained prior to accessing the survey. Participant confidentiality was maintained throughout this study by assigning code numbers to each pharmacist-participant, ensuring that personal identifiers were not linked to the data. All the data collected were stored securely in the researcher’s laptop, and backed up. Additionally, any published results were presented in their aggregate form, with no individual responses or identifying details disclosed, further safeguarding participant anonymity.

### Data Analysis

Quantitative data were entered into SPSS version 18.0 (SPSS Inc., Chicago, IL, USA) for descriptive analysis. Data were summarized using frequencies and percentages. The accuracy of the data was assessed by visual inspection by two data analysts who were independent of the study.

## RESULTS

### Demographics

A total of 12 pharmacists participated in the study, with the majority being female (66.7%). The largest age group was 20-29 years (41.7%), followed by 40-49 years (33.3%). Most participants had between 1 to 5 years of hospital experience (50%), and 58.3% held a BPharm as their highest qualification, while the remaining had postgraduate degrees (see Table 1).

**Table 1.** Demographics of Pharmacist-Respondents.

| **Variable** | **Category** | **Frequency (n=12)** | **Percentage (%)** |
| --- | --- | --- | --- |
| Sex | Female | 8 | 66.7 |
|  | Male | 4 | 33.3 |
| Age (years) | 20–29 | 5 | 41.7 |
|  | 30–39 | 2 | 16.7 |
|  | 40–49 | 4 | 33.3 |
|  | 50–59 | 1 | 8.3 |
| Experience (years) | <1 | 1 | 8.3 |
|  | 1–5 | 6 | 50.0 |
|  | 6–10 | 3 | 25.0 |
|  | 11–15 | 2 | 16.7 |
| Highest qualification | BPharm | 7 | 58.3 |
|  | MSc/PharmD | 5 | 41.7 |

### Frequency of pharmaceutical care intervention activities

Pharmacists in this study reported engaging in a variety of pharmaceutical care intervention activities (Figure 1), with the most common being therapeutic substitutions (87%) and providing drug information to physicians (83%). Other frequent activities included: preventing duplication of medication therapy and adjusting dosages. Less-commonly performed interventions involved counseling on drug storage and assisting with adverse drug reaction reporting (see Figure 1).

**Figure 1.**
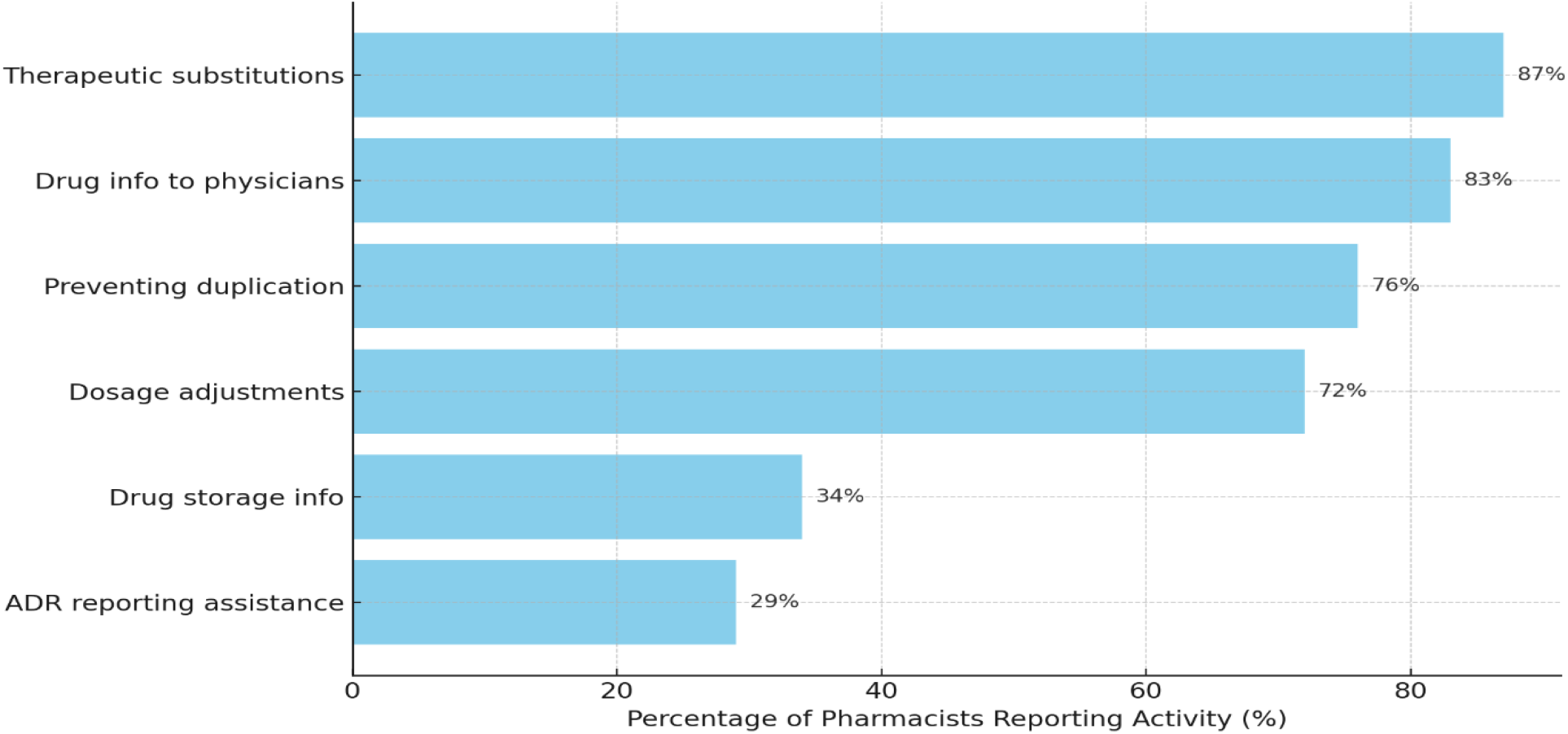
Frequency of Pharmacist Intervention Activities

### Frequency of reported barriers

Frequency of barriers to pharmaceutical care interventions as reported by the pharmacists in this study are displayed in Figure 2. The primary barriers to effective pharmaceutical care included: systemic and infrastructural challenges such as lack of access to patient records (92%), inadequate privacy (88%), and poor documentation systems (83%). Notably, most pharmacists did not perceive staffing levels or lack of clinical skills as significant limitations, reflecting strong professional engagement.

**Figure 2.**
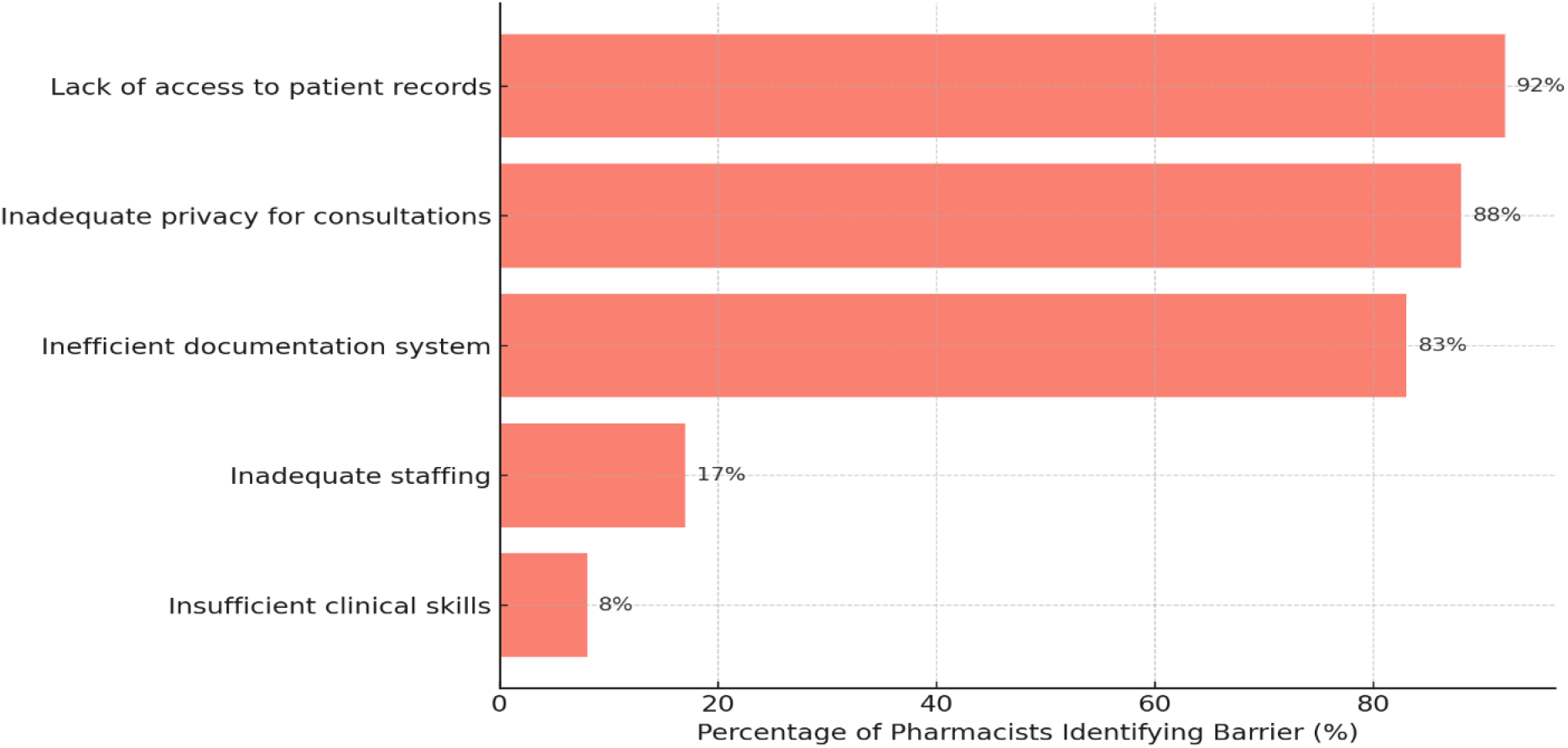
Frequency of Reported Barriers to Pharmacist Interventions

## DISCUSSION

Interpretation of the findings from this study are guided by Figure 1, which summarizes the frequency of pharmacists’ interventions; and Figure 2, which presents the pharmacist-reported barriers.

### Pharmaceutical care intervention activities

Pharmacists at this Nigerian secondary-care facility engaged in a range of clinically-valuable interventions (Figure 1), with therapeutic substitutions (87%), provision of drug information to prescribers (83%), and prevention of drug duplication (76%) being the most commonly reported activities. These practices are consistent with hospital pharmacy activities published in literature e.g. provision of drug information, making therapeutic substitutions, dosage adjustments (Langebrake et al. 2015), adverse drug reaction-reporting and prevention of duplicate therapy (Schatz & Weber, 2015) - reflecting pharmacists’ commitment to promoting patient safety and optimal therapy (Duarte et al., 2019). More recent studies suggest that these hospital pharmacy activities in some LMICs are not yet consistently common across facilities - and are reported as either still low in terms of scope, or not widely implemented; and are shaped by workforce and system-capacity constraints, with marked differences between hospitals and settings (Mwakawanga et al., 2025; Pathak et al., 2025). However, where they are actually implemented in LMICs (often in tertiary or specialty inpatient settings), pharmacists routinely make dose adjustments, therapeutic substitutions, prevent therapeutic duplication, and discontinue unnecessary therapy (Alsetohy et al., 2025). Global practice standards like the Basel Statements, continue to explicitly position hospital pharmacists as responsible for optimizing and monitoring medication use (Stepanovic et al., 2025); while national policy signals in some LMICs frame them as expected, even though uptake of these pharmacists’ activities remains uneven (Lambert et al., 2026).

### Barriers/Constraints to Pharmacists’ Interventions

In the current study, pharmaceutical care activities are hindered by significant structural barriers - most notably: lack of access to patient records (92%) and inadequate privacy for consultations (88%). These systemic issues are consistent with prior findings in West African and low-to middle-income country (LMIC) contexts (Hamza et al., 2025; Sendekie et al., 2025). Another major barrier to effective pharmaceutical care as reported by pharmacists in this study, was inefficient documentation systems (83%), with pharmacists reporting lack of standardized documentation practices.

### Limited access to patient records

Despite the high frequency of intervention activities shown in Figure 1, lack of access to comprehensive patient records emerged as the most significant barrier (Figure 2). Restricted access to medical histories, laboratory results, and diagnostic information limits pharmacists’ ability to perform comprehensive medication reviews, monitor therapy, and flag contraindications or drug interactions. Similar challenges have been reported where fragmented or inaccessible health records limit the effectiveness of pharmacist-led interventions (Bronkhorst et al., 2020; Hamza et al., 2025). It has been reported that in hospitals without electronic medical records (EMRs), communication between pharmacists and prescribers is often delayed or incomplete, leading to “misaligned processes of care (Mercer et al., 2018).

### Privacy limitations

Inadequate privacy for patient consultations was reported by the majority of pharmacists (Figure 2) and corresponds with the lower frequency of patient-directed interventions (Figure 1). The need for private spaces for patient counseling is critical, and the absence of such environments not only breaches patient confidentiality but can reduce the willingness of patients to disclose sensitive information such as medication non-adherence, mental health challenges, or sexual and reproductive health concerns. This was similarly observed in other studies from Nigeria, and from one review of global experiences, where pharmacists highlighted privacy limitations as a key factor reducing the impact of their counseling efforts (Ipingbemi et al., 2023; Chiedu & Collins, 2011; Homayounifar, et al., 2025).

### Documentation challenges

Inefficient documentation systems were identified as a major barrier (Figure 2). Documentation is essential for continuity of care, interprofessional communication, and quality improvement. Without standardized mechanisms for recording interventions, pharmacists’ contributions may not be consistently integrated into patient care plans or institutional learning processes. In the present study, the high frequency of interventions (shown in Figure 1) may therefore underestimate pharmacists’ actual clinical impact, as undocumented activities are less likely to influence future care. Without standardized documentation systems, pharmacists have reportedly struggled with effectively recording and communicating their interventions – which in turn undermines interdisciplinary collaboration and diminishes the visibility of pharmacist contributions. In general, improvements in electronic documentation platforms have been associated with enhanced care-tracking and pharmacist accountability (Singer & Duarte Fernandez, 2015; Falevai & Hassandoust, 2025).

### Professional capacity within constrained systems

Notably, pharmacists in this study did not identify inadequate staffing or insufficient clinical knowledge as major barriers in Figure 2. When interpreted alongside the frequent and somewhat time-involving interventions shown in Figure 1, this finding suggests a professionally-capable and adequate workforce operating within organizational and infrastructural constraints. This distinction underscores the importance of system-level reforms to enable pharmacists to fully apply their clinical expertise. Policy makers and professional associations in low-middle-income countries (LMICs) like Nigeria could further support these needs by developing national guidelines for pharmacist documentation, expanding electronic medical record (EMR) access, and incentivizing infrastructure improvements. For instance, evidence from Egypt demonstrates that integrating clinical pharmacists with electronic medical record systems and medication management tools improves their ability to detect and resolve drug therapy problems, resulting in significant clinical and economic benefits (Alsetohy et al., 2025). While pharmacists’ interventions in LMICs have been shown to significantly improve clinical and economic outcomes (Okoro & Nduaguba, 2021), experts emphasize that establishing formal clinical positions, adequate infrastructure, and supportive regulatory frameworks are still needed to fully strengthen pharmacy practice across LMICs (Babar, 2021).

## LIMITATIONS

This study was conducted in a single facility with a small sample size, which limits its generalizability. Secondly, the findings relied on self-reported data and may be subject to recall or social desirability bias. Also, the cross-sectional design precludes causal inference between barriers and frequency of pharmacists’ interventions. Last of all, patients’ clinical outcomes and patient-reported experiences with pharmaceutical care were not assessed.

## RECOMMENDATIONS

Based on this study, in order to enhance pharmaceutical care delivery in LMICs with similar settings, the following actions are recommended: (1) improve pharmacists’ access to comprehensive patient records to support medication management or reviews, as well as and collaborative care; (2) provide private or semi-private consultation spaces to enable private discussions about medication concerns and health conditions; and (3) implement standardized documentation tools to record pharmacist interventions and strengthen continuity of care and interprofessional communication.

## CONCLUSION

The pharmacists in this secondary-care facility reported frequent engagement in clinically meaningful interventions that support medication optimization and patient safety. However, the delivery of comprehensive pharmaceutical care is constrained by system-level barriers, particularly limited access to patient records, inadequate privacy for consultations, and inefficient documentation systems. Addressing these barriers is essential to strengthen patient-centered pharmaceutical care in secondary-care hospital environments.

## Data Availability

All data produced in the present study are available upon reasonable request to the authors

## REFERENCES

Akande-Sholabi, W., & Akinbitan, A. (2022). Assessment of attitude, practice and barriers to pharmaceutical care among community pharmacists in Ibadan. Nigerian Journal of Pharmaceutical Research, 18(1), 75–84. 10.4314/njpr.v18i1.8

Alsetohy, W. M., El-Fass, K. A., El Hadidi, S., Zaitoun, M. F., Badary, O., Ali, K. A., … & Zaki, H. V. (2025). Economic impact and clinical benefits of clinical pharmacy interventions: A six-year multi-center study using an innovative medication management tool. PloS one, 20(1), e0311707. 10.1371/journal.pone.0311707

Babar, Z. U. D. (2021). Ten recommendations to improve pharmacy practice in low and middle-income countries (LMICs). Journal of Pharmaceutical Policy and Practice, 14(1). 10.1186/s40545-020-00288-2

Bond, C. A., Raehl, C. L., & Franke, T. (2002). Clinical pharmacy services, hospital pharmacy staffing, and medication errors in United States hospitals. Pharmacotherapy: The Journal of Human Pharmacology and Drug Therapy, 22(2), 134–147.10.1592/phco.22.3.134.33551

Duarte, N. C., Barbosa, C. R., Tavares, M. G., Dias, L. P., Souza, R. N., & Moriel, P. (2019). Clinical oncology pharmacist: effective contribution to patient safety. Journal of oncology pharmacy practice, 25(7), 1665–1674. 10.1177/1078155218807748

Homayounifar, F., Khosravi, M., Davar, G., Negahdaripour, M., & Joulaei, H. (2025). Implications Derived from Global Experiences of Barriers to Integrating Community Pharmacists into Primary Healthcare: A Scoping Review. Journal of Primary Care & Community Health, 16, 21501319251371825. 10.1177/215013192513718

Bronkhorst, E., Gous, A. G., & Schellack, N. (2020). Practice guidelines for clinical pharmacists in middle to low income countries. Frontiers in Pharmacology, 11, 978. 10.3389/fphar.2020.00978

Chiedu, A. K., & Collins, O. (2011). Attitude and practice of community pharmacists towards pharmaceutical care in Nigeria. 10.22270/jddt.v9i6-s.3786

Cipolle, R. J., Strand, L. M., & Morley, P. C. (2012). Pharmaceutical care practice: the patient-centered approach to medication management services. Manag. Serv, 1, 20.

Falevai, I., & Hassandoust, F. (2025). Enhancing Transplantation Care with eHealth: Benefits, Challenges, and Key Considerations for the Future. Future Internet, 17(4), 177. 10.3390/fi17040177

Hamza, M.A., Imran, F., Ahmed, A. et al. Medication therapy management in Pakistan: a cross-sectional evaluation of pharmacists’ knowledge attitudes, practices, and barriers. J Pharm Health Care Sci 11, 85 (2025). 10.1186/s40780-025-00493-8

Hepler, C. D., & Strand, L. M. (1990). Opportunities and responsibilities in pharmaceutical care. American journal of hospital pharmacy, 47(3), 533–543. 10.1093/ajhp/47.3.533

Ipingbemi, A. E., Ajanaku, O. O., & Umaru, O. T. (2023). Assessment of community pharmacists’ knowledge and counselling practices on oral contraceptives use. *Journal of Contemporary Studies in Epidemiology and Public Health, 4*(2), ep23008. 10.29333/jconseph/13848

Kaboli, P. J., Hoth, A. B., McClimon, B. J., & Schnipper, J. L. (2006). Clinical pharmacists and inpatient medical care: A systematic review. *Archives of Internal Medicine, 166*(9), 955–964. doi:10.1001/archinte.166.9.955

Lambert, M., Mmassy, J., Kabissi, S., Benizeth, C., Baltazary, G., Maganda, B. A., & Taxis, K. (2026). Lessons from the field: implementing pharmacy services in the Tanzanian hospital setting. International Health, 18(1), 1–4. 10.1093/inthealth/ihaf079

Langebrake, C., IhbelJHeffinger, A., Leichenberg, K., Kaden, S., Kunkel, M., Lueb, M., … & Hohmann, C. (2015). Nationwide Evaluation of DaylJtolJDay Clinical Pharmacists’ Interventions in G erman Hospitals. Pharmacotherapy: The Journal of Human Pharmacology and Drug Therapy, 35(4), 370–379. 10.1002/phar.1578

Lee, M., Badowski, M. E., Acquisto, N. M., Covey, D. F., Fox, B. D., Gaffney, S. M., … & Turner, K. (2017). ACCP template for evaluating a clinical pharmacist. Pharmacotherapy: The Journal of Human Pharmacology and Drug Therapy, 37(5), e21–e29. 10.1002/phar.1927

Mercer, K., Burns, C., Guirguis, L., Chin, J., Dogba, M. J., Dolovich, L., … & Grindrod, K. A. (2018). Physician and pharmacist medication decision-making in the time of electronic health records: mixed-methods study. JMIR Human Factors, 5(3), e9891. doi:10.2196/humanfactors.9891

Mwakawanga DL, Mutagonda RF, Mlyuka HJ, Mikomangwa WP, Kilonzi M, Kibanga WA, et al. Improving the provision of clinical pharmacy services in low- and middle-income countries: a qualitative study in tertiary health facilities in Tanzania. BMJ Public Health. 2025;3:e001776. 10.1136/bmjph-2024-001776

Ogbonna, B. O., Ezenduka, C. C., Soni, J. S., & Oparah, A. C. (2015). Limitations to the dynamics of pharmaceutical care practice among community pharmacists in Enugu urban, southeast Nigeria. Integrated Pharmacy Research and Practice, 49–55.10.2147/IPRP.S82911

Okoro, R. N., & Nduaguba, S. O. (2021). Community pharmacists on the frontline in the chronic disease management: The need for primary healthcare policy reforms in low and middle income countries. Exploratory Research in Clinical and Social Pharmacy, 2, 100011. 10.1016/j.rcsop.2021.100011

Oparah, A.C., Eferakeya, A.E. Attitudes of Nigerian Pharmacists towards Pharmaceutical Care. Pharm World Sci 27, 208–214 (2005). 10.1007/PL00022056

Pathak, N., Barma, S., Jha, P. K., Shrestha, P., Yadav, U., Sah, A. K., … & Shrestha, S. (2025). Strengthening Hospital Pharmacy Practice in Nepal Through the Minimum Service Standards Checklist. Integrated Pharmacy Research and Practice, 149–157. 10.2147/IPRP.S557016

Schatz, S., & Weber, R. J. (2015). Adverse drug reactions. Pharmacy Practice, 1(1), 16–23. https://www.accp.com/docs/bookstore/psap/2015B2.SampleChapter.pdf?fbclid=IwAR0y2CBAJV-RqmiaTN7vYAQxsIIsdCIBh16ZejTJfghAui2JfJ3U4T9zZcg. Accessed on 01/23/2026

Sendekie, A. K., Belachew, E. A., Limenh, L. W., Chanie, G. S., Bizuneh, G. K., Dagnaw, A. D., … & Abate, B. B. (2025). Roles and barriers of community pharmacy professionals in the prevention and management of noncommunicable diseases in Ethiopia: a systematic review. Frontiers in Public Health, 13, 1485327. 10.3389/fpubh.2025.1485327

Singer, A., Duarte Fernandez, R. The effect of electronic medical record system use on communication between pharmacists and prescribers. BMC Fam Pract 16, 155 (2015). 10.1186/s12875-015-0378-7

Stepanovic, M., Dabliz, R., Moss, R. J., Moles, R. J., Vaillancourt, R., Penm, J., & Eckel, S. F. (2025). The 2024 updated Basel Statements on the future of hospital pharmacy. American Journal of Health-System Pharmacy, zxaf131. 10.1093/ajhp/zxaf131

